# How safe is Covishield (ChAdOx1nCoV-19) vaccine? Experience from a tertiary care hospital in South India

**DOI:** 10.1101/2021.03.16.21253744

**Authors:** Leeberk Raja Inbaraj, Carolin Elizabeth George, Nirupama Navamani Franklyn

## Abstract

**Background:** COVID vaccine has been recently rolled out for Health care workers in India. Though vaccine trials and data are available, people, including HCWs, need more real-life data from their contexts to get over the vaccine hesitancy. We aimed to determine the incidence of Adverse Event Following Immunisation (AEFI) among HCWs after their first dose of ChAdOx1nCoV-19 vaccine

**Methods:** We conducted a cross-sectional study among 218 staff of a tertiary care hospital. We circulated a google form with informed consent on the hospital WhatsApp platform and asked them to self-report their vaccination experience.

**Results:** Two thirds (69.7% (152/218), 95% CI: 63.1-75.7) had minor AEFI, and none had severe AEFI. Body ache (46.8% (102/218), 95% CI :40 – 53.6) was the most common symptom followed by headache (30.3% (66/218), 95% CI :24.2 – 36.8) and fever (22% 948/218), 95% CI :16.7 – 28.1). Most of them (39.4% (87/218) 95% CI:32.9 – 46.2) experienced symptoms within 4-24 hours after taking the vaccine while 22.3% (49/218) (95% CI: 17.1 – 28.6) experienced it after a day. Majority of the HCWs (78.9% (172/218), 95% CI: 72.8 – 84.1) were anxious before the vaccination. Younger age group and female gender were significantly associated with AEFI compared to their counterparts.

**Conclusion:** HCWs experienced minor, self-limiting AEFI with the first dose of ChAdOx1nCoV-19. The hospital reported no serious AEFI following the vaccination

## Introduction

Ever since the Corona Virus Disease (COVID-19) pandemic, India has recorded more than 10.5 million cases and 150000 deaths.^1^ India began its vaccine roll out against COVID-19 on16 January 2021 to Healthcare Workers (HCWs), almost a year after the index case was detected in the sub-continent. The country has approved two vaccines so far. Covaxin, a whole-Virion Inactivated VeroCell vaccine developed by Bharat Biotech in collaboration with Indian Council of Medical Research (ICMR) andCovishield (ChAdOx1nCoV-19), recombinant vaccine manufactured by SerumInstitute of India in partnership with Oxford-AstraZeneca.^2^ The Government of India (GOI) has procured 11 and 5.5 million doses of Covisheild and ChAdOx1nCoV-19, respectively for the initial rollout. ^3^

Though the country has experienced the pandemic’s fury, many rumours were associated with the vaccine, even among the Health Care workers (HCWs). Hence vaccine was welcomed with the paradox of urgency and hesitancy. Though vaccine trials and data are available, people, including HCWs, need more real-life data from their contexts to get over the vaccine hesitancy. We aimed to determine the incidence of Adverse Event Following Immunisation (AEFI) among HCWs after their first dose of ChAdOx1nCoV-19 vaccine.

## Methodology

We conducted a cross-sectional study among HCWs of Bangalore Baptist Hospital (BBH), 350-bed tertiary care teaching hospital with close to 1500 staff located in South India. The hospital had treated more than 2000 COVID-19 in-patients in the past ten months. Out of 1376 staff members registered for the vaccine, only 758 (55.08%) took the vaccine.

Assuming 50% of the vaccinated HCWs develop minor AEFI, a minimum sample size of 97 was calculated with a relative precision of 20%. We circulated a google form with informed consent on the hospital WhatsApp platform and asked them to self-report their vaccination experience. The questionnaire contained demographic information, their anxiety level before taking the vaccine and questions on AEFI. Data were analyzed in Statistical Package for the Social Sciences (SPSS) version 20.0. Socio-demographic factors, history of COVID-19 infection, comorbid conditions, level of anxiety were dichotomized and tested for significance using the chi-square test.

## Results

Out of 758 vaccinated HCWs, 218 (28.6%), have responded to the survey (Table.1). The mean age of the participants was 36 years (SD-9.5). Diabetes (19 (8.7% (19/218) 95% CI : 5.3-13.2), and hypertension (8.2% (18/218), 95% CI : 3.5-10.5) were the most common comorbidities.

**Table.1.**
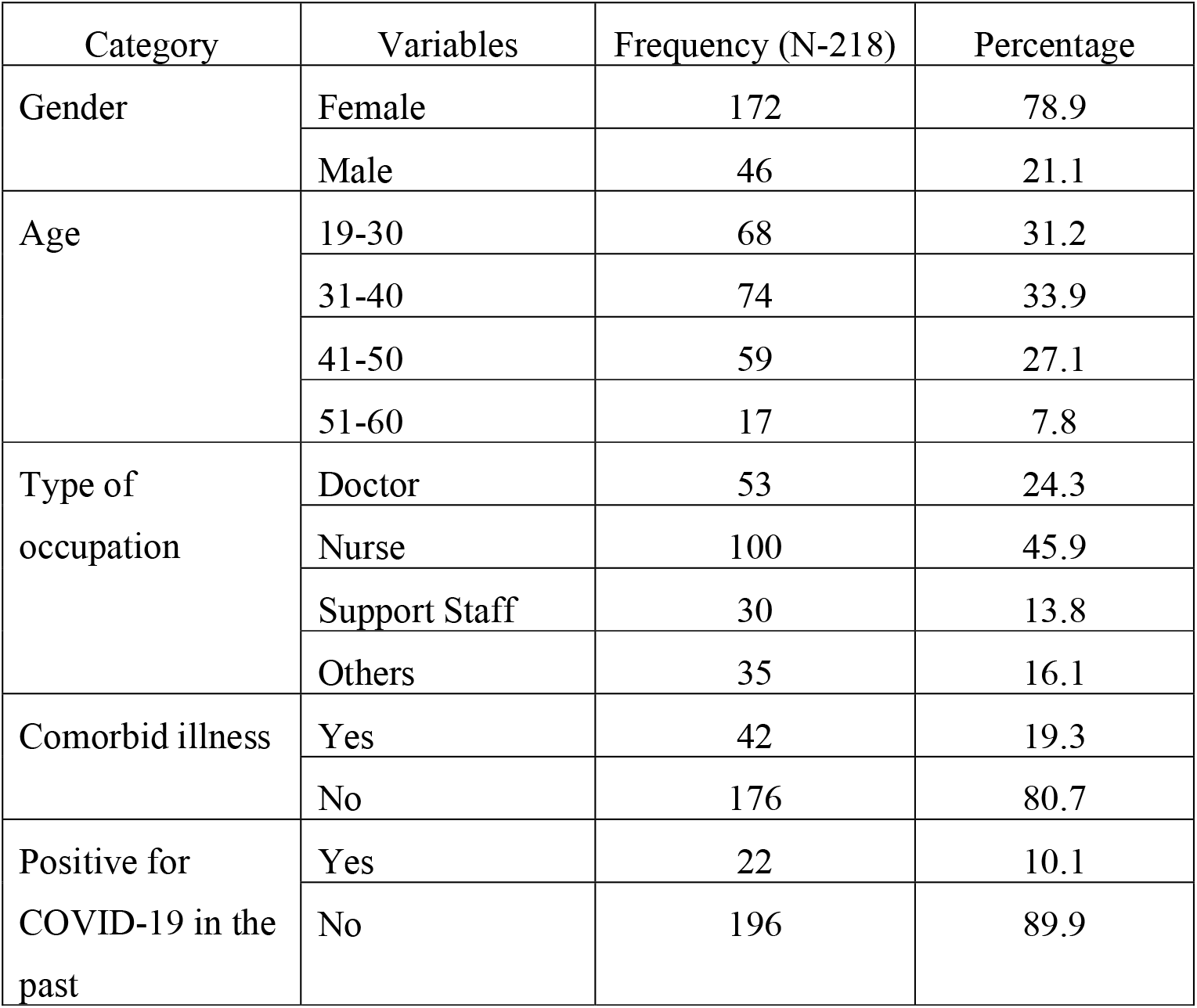
Sociodemographic characters of the study population.

Almost two thirds (69.7% (152/218), 95% CI: 63.1-75.7) had minor AEFI, and none had severe AEFI. When HCWs were asked to rate the severity of AEFI, more than half of them (51.8% (113/218), 95% CI: 44.9-58.6) perceived it as mild, while (3.2% (7/218), 95% CI:1.3 – 6.5) rated it as severe. Body ache (46.8% (102/218), 95% CI: 40-53.6) was the most common symptom followed by headache (30.3% (66/218), 95% CI:24.2-36.8) and fever (22% (48/218), 95% CI: 16.7-28.1) (Table.2). Many (39.9%, 95% CI:33.3 – 46.7) took medications for their symptoms, and one required admission for observation. Most of them (39.4% (87/218) 95% CI: 32.9-46.2) experienced symptoms within 4-24 hours after taking the vaccine while 49 (22.3% 95% CI:17.1 – 28.6) experienced it after a day.

**Table.2.** Self-reported AEFI by the study population.

| **Symptoms** | **Frequency** | **Percentage** | **95% CI** |
| --- | --- | --- | --- |
| **Local symptoms** |  |  |  |
| Pain | 166 | 76.1 | 69.9 – 81.6 |
| Swelling | 36 | 16.5 | 11.8 – 22.1 |
| Induration | 8 | 3.7 | 1.6 – 7.1 |
| **Symptoms related to the injection arm** |  |  |  |
| Stiffness of injection arm | 25 | 11.5 | 7.5 – 16.4 |
| Weakness of injection arm | 28 | 12.8 | 8.7 – 18 |
| Numbness of injection arm | 8 | 3.7 | 1.6 – 7.1 |
| **Systemic symptoms** |  |  |  |
| Headache | 66 | 30.3 | 24.2 – 36.8 |
| Fever | 48 | 22 | 16.7 – 28.1 |
| Body ache | 102 | 46.8 | 40 – 53.6 |
| Felt weak | 68 | 31.2 | 25.1 – 37.8 |
| **GI symptoms** |  |  |  |
| Vomiting | 8 | 3.7 | 1.6 – 7.1 |
| Abdominal pain | 9 | 4.1 | 1.9 – 7.6 |
| Loose stools | 5 | 2.3 | 0.75 – 5.2 |
| **Allergic symptoms** | 5 | 2.3 | 0.75 – 5.2 |

Majority of the respondents (78.9% (172/218), 95% CI:72.8 – 84.1) were anxious before the vaccination. Anxiety prior to the vaccination was reported as mild by (50.9 (111/218) 95% CI:44 – 57.7) and as severe by 8.7% (19/218) (95% CI 5.3 – 13.2). More than two-thirds (61.5% (134/218), 95% CI:54.6 – 67.9) felt happy to have received the vaccine, and 88.1% (192/218) (95% CI:83 – 92) said they would be taking the next dose of vaccine.

Level of anxiety, type of occupation, comorbid conditions, history of COVID infection in the past was not significantly associated with AEFI. Younger age group (23.9%Vs 6.6%) and female gender (20.9% Vs 6.5%) were significantly associated with AEFI compared to their counterparts (p<0.05).

## Discussion

We found that ChAdOx1nCoV-19 has a good safety profile. Our findings concur with phase 2-3 safety trial data ofChAdOx1nCoV-19where 88% of the participants between the age of 18 and 55 years experienced at least one local symptom.^4^ After rolling out 45 lakh doses, the GOI reported an overall AEFI of 0.18%, of which 0.0007% required hospitalizations. ^5^ Most of these were ‘injection-related side effects’ and ‘immunization anxiety-related reaction similar to our study. Nineteen vaccinated HCW’s have died in the country so far, but none have been attributed to the vaccine.

High prevalence of anxiety (78.9%) is not surprising owing to various reasons. Fast-tracked vaccine development, limited knowledge about long term effects, controversies on Covaxin’sapproval without phase-3 trial, and social media could have mounted the anxiety.^6^ Many still believe that vaccines are still a ‘hit or miss strategy’ than an adequately evaluated prevention measure.Even in a hospital, where the risk of infection is high, the percentage of vaccine takers was only 55.08 %. Anxiety can be one of the main reasons for vaccination hesitancy in our centre and in the country.^7^ However, we cannot comment on this, as our sample had only HCWs who took the vaccine.

In our study, younger age group and female gender were associated with minor AEFIs. High reactogenicity of ChAdOx1nCoV-19 in the younger age group was documented.^4^ However, we did not find any supporting evidence for high reactogenicity among women, and this may be due to the higher proportion of women in our sample. Though anxiety is thought to be associated with AEFI, we could not find this association in our study. There is also a possibility that most people with severe anxiety would have refrained from vaccination. ^8^

To the best of our knowledge, this is the first study from the country exploring the incidence of AEFI among HCW. The study has an adequate sample size and a reasonable response rate. The study used a WhatsApp platform; hence we would have missed those who did not have smartphones.

## Conclusion

HCWs experienced minor, self-limiting AEFI with the first dose of ChAdOx1nCoV-19. The hospital reported no serious AEFI following the vaccination. ChAdOx1nCoV-19 has a good safety profile, and we believe our findings will encourage people to come forward and receive this public health tool to break the chain of transmission.

## Data Availability

Data is available with the corresponding author at request

## Declaration

### Ethics approval and consent to participate

This study was approved by Institutional Review Board of Bangalore Baptist Hospital. An electronic informed consent was taken from all the participants of the study

### Availability of data and materials

The datasets used and/or analyzed during the current study are available from the corresponding author on reasonable request.

### Competing interests

None

### Funding

None

## Authors’ contributions

LR conceived the idea, developing the study instrument, conducted analysis, interpreted data,prepared the manuscript. GE conceived the idea of the study, interpreted the data and contributed in writing of the manuscript. NF conceptualized and assisted with design of the study, tool developmentand reviewed the manuscript. All authors read and reviewed the final manuscript.

## Acknowledgments

We extend our thanks to staff of Bangalore Baptist Hospital in taking part in this study. We would like to extend our gratitude to Ms.Sherin Manichen for her assistance in analysis

## Notes

### Competing Interest Statement

The authors have declared no competing interest.

### Funding Statement

No external funding was received

### Author Declarations

The study was approved by Institutional Review Board of Bangalore Baptist Hospital

